# Diagnostic accuracy of Loop mediated isothermal amplification coupled to Nanopore sequencing (LamPORE) for the detection of SARS-CoV-2 infection at scale in symptomatic and asymptomatic populations

**DOI:** 10.1101/2020.12.15.20247031

**Authors:** Anetta Ptasinska, Celina Whalley, Andrew Bosworth, Charlotte Poxon, Claire Bryer, Nicholas Machin, Seden Grippon, Emma L Wise, Bryony Armson, Emma L A Howson, Alice Goring, Gemma Snell, Jade Forster, Chris Mattocks, Sarah Frampton, Rebecca Anderson, David Cleary, Joe Parker, Konstantinos Boukas, Nichola Graham, Doriana Cellura, Emma Garratt, Rachel Skilton, Hana Sheldon, Alla Collins, Nusreen Ahmad, Simon Friar, Tim Williams, Keith M Godfrey, Zandra Deans, Angela Douglas, Sue Hill, Michael Kidd, Deborah Porter, Stephen P Kidd, Nicholas J Cortes, Veronica Fowler, Tony Williams, Alex Richter, Andrew D Beggs

## Abstract

**Introduction:** Rapid, high throughput diagnostics are a valuable tool, allowing the detection of SARS-CoV-2 in populations, in order to identify and isolate people with asymptomatic and symptomatic infections. Reagent shortages and restricted access to high throughput testing solutions have limited the effectiveness of conventional assays such as reverse transcriptase quantitative PCR (RT-qPCR), particularly throughout the first months of the COVID-19 pandemic. We investigated the use of LamPORE, where loop mediated isothermal amplification (LAMP) is coupled to nanopore sequencing technology, for the detection of SARS-CoV-2 in symptomatic and asymptomatic populations.

**Methods:** In an asymptomatic prospective cohort, for three weeks in September 2020 health care workers across four sites (Birmingham, Southampton, Basingstoke and Manchester) self-swabbed with nasopharyngeal swabs weekly and supplied a saliva specimen daily. These samples were tested for SARS-CoV-2 RNA using the Oxford Nanopore LamPORE system and a reference RT-qPCR assay on extracted sample RNA. A second retrospective cohort of 848 patients with influenza like illness from March 2020 – June 2020, were similarly tested from nasopharyngeal swabs.

**Results:** In the asymptomatic cohort a total of 1200 participants supplied 23,427 samples (3,966 swab, 19,461 saliva) over a three-week period. The incidence of SARS-CoV-2 detection using LamPORE was 0.95%. Diagnostic sensitivity and specificity of LamPORE was >99.5% in both swab and saliva asymptomatic samples when compared to the reference RT-qPCR test. In the retrospective symptomatic cohort, the incidence was 13.4% and the sensitivity and specificity were 100%.

**Conclusions:** LamPORE is a highly accurate methodology for the detection of SARS-CoV-2 in both symptomatic and asymptomatic population settings and can be used as an alternative to RT-qPCR.

## Introduction

COVID-19, caused by an emergent novel *betacoronavirus* known as SARS-CoV-2; represents a public health emergency. First identified in Wuhan, China(1) in December 2019 it has spread rapidly across the world, causing, at the time of writing, over 700,000 deaths with 19.4 million cases detected.

Rapid detection of infected cases in order to limit transmission, remains challenging as most validated methods utilise reverse transcription quantitative polymerase chain reaction (RT-qPCR) (2) targeting one or more gene targets for SARS-CoV-2. Although considered the gold standard for diagnosis, RT-qPCR is laborious can be difficult to scale up for mass-testing, and competition for reagents/equipment from many laboratories may lead to widespread reagent shortages. Initially in the outbreak, labs throughout the United Kingdom utilised primers that were designed to target sequences within the RNA dependent polymerase gene (RdRp) (3), however these lacked sensitivity (4). RT-qPCR Primer sets were introduced that targeted the envelope (*E*), nucleocapsid (*N*) and *ORF1ab genes*, which provided necessary increased sensitivity (5). As all these assays depend on RT-qPCR technology, if viral gene targets are amplified singly for a sample within a series of wells, this reduces the overall capacity of RT-qPCR. Multiplexing of gene targets (6) using several fluorescent dyes within the same reaction volume increases capacity, but the majority of machines in common use can only utilise a maximum of 4-5 dyes (limiting to 4-5 gene targets) and also process between 96-384 samples simultaneously. A typical RT-qPCR reaction requires approximately 60-90 minutes to process, meaning that these are rate limiting steps, impeding the ability to expand capacity to allow rapid and widespread detection of cases.

Loop mediated isothermal amplification (LAMP) offers an alternative to RT-qPCR (8). It utilises a strand displacement, replicating DNA polymerase utilising multiple primer sets (and specific “Loop primers” to increase specificity) to rapidly amplify DNA using a continuous temperature. This reaction typically takes 20-30 minutes which is considerably quicker than PCR. The LAMP reaction can also be coupled to a reverse transcriptase, allowing detection of RNA (RT-LAMP) (9)

Nanopore sequencing allows rapid sequencing using protein nanopores embedded in a lipid membrane. Electrical current passes through the nanopores and changes in electrical conductivity occur as molecules of nucleic acid translocate through the pores. Each base produces a characteristic change in current which is detected and converted into a readable trace (10). This allows the sequencing data to be analysed in real time, and the process can be halted once sufficient data is recorded. Nanopore sequencing technology allows all the advantages of conventional next generation sequencing technology, especially the capacity to perform very high (limited only by the number of barcodes available) sample multiplexing within the same sequencing run.

LamPORE is a combination of LAMP and Nanopore sequencing, developed by Oxford Nanopore (11) that utilises barcoded LAMP reactions to generate amplicons from SARS-CoV-2 viral RNA. These can then be multiplexed via sample barcoding and pooled onto a flow cell for sequencing. This technology has a theoretical maximum capacity of 15,000 samples per GridIon Mk 1 machine (Oxford, Nanopore) per 24 hours, allowing scalability and high throughput.

Use of alternative sampling strategies, such as saliva, could theoretically increase capacity over swabbing (because of less pressure on the supply chain) and increase compliance because of the less invasive nature of sampling of saliva. There is also evidence that viral loads in saliva are high prior to symptoms and early in infection (7).

This study aimed to assess the assay performance characteristics of the LamPORE SARS-CoV-2 Detection Assay against the gold standard RT-qPCR for SARS-CoV-2 detection in both symptomatic and asymptomatic populations from multiple independent centres.

## Methods

### Study design

The study consisted of a retrospective and prospective diagnostic accuracy study comparing the performance of LamPORE sequencing of the *ORF1ab, N2* and *E* gene targets of SARS-CoV-2 against RT-qPCR of the *ORF1ab* and *N1* gene targets of SARS-CoV-2 (12).

### Participants

For the prospective study 1200 health care workers at high risk of asymptomatic transmission were recruited as part of a consented National Health Service England and NHS Improvement (NHSE/I) service evaluation in September 2020. Prospective participants performed naso-pharyngeal self-swabs at day 0, 7, 14 and 21 as well as daily saliva sampling for 21 days (Figure 1). Any symptoms were reported in a symptom diary. Participants were recruited from staff working within five sites: University Hospitals Birmingham NHS Foundation Trust, Birmingham Women’s’ and Children’s’ NHS Foundation Trust, University Hospital Southampton NHS Foundation Trust, Hampshire Hospitals NHS Foundation Trust and Manchester University NHS Foundation Trust. The retrospective study was undertaken by collating surplus sample from patients having diagnostic samples sent to the Public Health England West Midlands laboratory for respiratory panel testing for influenza like illness (ILI) from January 2020 – June 2020 (National Research Ethics Service Committee West Midlands - South Birmingham 2002/201 Amendment Number 4).

**Figure 1:**
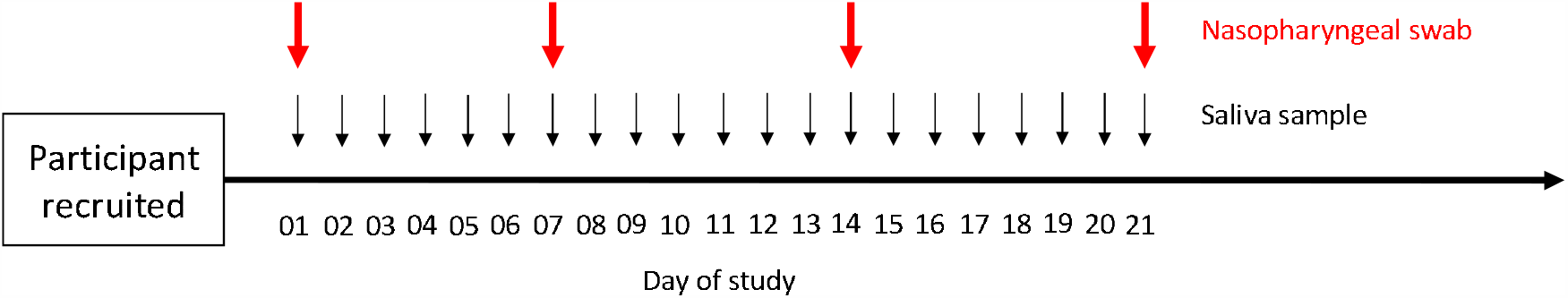
Graphical representation of recruitment strategy for collecting saliva and swabs. Day of study is shown below thick black horizontal line. Nasopharyngeal swab sampling timings are represented by thick red vertical arrows. Saliva sample timings represented by thin black vertical arrows.

### Test methods

#### Sampling

For swab-based tests, participants underwent self-directed nasopharyngeal swabbing using flocked swabs containing viral transport medium. For saliva-based tests participants were instructed to dribble at least 1ml of saliva into a universal specimen container without any additive. Samples were tested immediately if returned on the day of testing or if received on Friday then stored for a maximum of 4 days at 4°C then tested.

### Clinical material used for analytical performance

To assess the limit of detection, a tenfold? dilution series (from 20,000 copies/ml to 0.2 copies/ml) of droplet digital polymerase chain reaction (ddPCR) quantified SARS-CoV-2 was constructed from the lysate of the Public Health England SARS-CoV-2 reference strain grown on Vero E6 cells and diluted in nuclease free water. This was tested by RT-qPCR and LamPORE in triplicate. For intra- and inter-assay precision, and reproducibility, 20,000 copies/ml of the same extracted SARS-CoV-2 RNA from the prepared dilution series was used. A panel of respiratory viruses (Zeptometrix Respiratory Panel R2, Buffalo NY 14202) was used to assess specificity of the LamPORE assay.

#### RNA extraction

For all samples at the Birmingham site, either viral transport medium (VTM) or saliva was supplied in screw capped tubes and heated to 56°C for a minimum of 30 minutes in order to inactivate live SARS-CoV-2 virus. The specimen was then aliquoted into a deep well sample plate, then 600uL of VTM or saliva was transferred to an extraction plate where RNA extraction was performed either via bead based or silica column methodology (depending on site) into an elution volume of 200uL. Extracted RNA was stored at −80C until required, with a minimum of freeze thaw cycles. For every batch of RNA extraction performed (95 samples/batch) an RNA extraction control was utilised consisting of heat inactivated SARS-CoV-2 virus grown on Vero-E6 cells (PHE SARS-CoV-2 England reference strain) normalised to 20,000 copies/ml via droplet digital PCR targeted against the E (Envelope) gene of SARS-CoV-2. At HHFT site, RNA extraction was performed on 200 µL VTM or saliva using the Maxwell® RSC Viral Total Nucleic Acid Purification Kit (Promega UK Ltd., Southampton, UK) on a Maxwell® RSC 48 Instrument (Promega UK Ltd., Southampton, UK), according to the manufacturer’s guidelines. Prior to loading onto the instrument, samples added to lysis buffer were inactivated at room temperature and then at 56°C for 10 minutes each. RNA was eluted in 50 µL of nuclease free water.

#### Reference test (RT-QPCR)

Single step reverse transcription quantitative polymerase chain reaction (RT-QPCR) against the ORF1ab and N1 gene targets of SARS-CoV-2 was carried out using the CerTest ViaSure SARS-CoV-2 real time PCR kit (CerTest Biotech SL, Zaragoza, Spain) according to manufacturer’s instructions for use (IFU) on ThermoFisher QuantStudio 5 or BioMolecular Systems MIC instruments, using 5uL of extracted RNA per reaction. The Reference test has been previously verified against gold standard RT-qPCR assays recommended by the WHO (2, 12)

#### Comparator test (LamPORE)

For each sample, initially 5uL of LAMP primer mix (LPM) was mixed with 25uL LAMP master mix (LMM) in each well of a 96 well plate, then 20uL of RNA sample was added and mixed thoroughly with a pipette. A positive control (CTL) and negative control (NTC) was added to each plate before sealing with a foil seal. The plate was then incubated at 65°C for 30 mins, then at 80°C for 5 minutes on a thermal cycler. Column based plate pooling of LAMP product was then carried out by combining 2uL of LAMP products from each well of a column into a 0.2uL PCR tube. Rapid sample barcoding was then carried out using a modified ONT-RSB (Rapid Sample Barcoding) protocol and all barcoded sequencing libraries columns were purified and pooled into a single tube which was cleaned according to manufacturer’s instructions using magnetic beads and run on an OND-FLO-M106D MinIon R.9.4.1 flow cell for a 1 hour run. Run monitoring was carried out using MinKnow and a proprietary Guppy/VSEARCH/SnakeMake pipeline that aligned reads to the viral target genes and a human beta-actin (*ACTB*) internal control and reported results in absolute reads per sample per gene.

### Analysis

#### Sample size calculation

Sample size was determined pragmatically, based on the incidence seen in the United Kingdom at the time of the study (1%). Sample size was calculated using R code (using R 3.6.3 (13)) from the methodology of Stark et al (14) for binary diagnostic test outcomes (a=0.05,b=0.90) setting a base sensitivity and specificity of RT-qPCR of 95% and 99% respectively. We aimed to be able to detect a change of sensitivity & specificity of 10% in LamPORE respectively giving a sample size of greater than 9,600 in the prospective cohort.

#### Results interpretation

The readers of RT-qPCR and LamPORE tests were blinded to any clinical information relating to study participants. For the RT-qPCR reference assay, SARS-CoV-2 was said to be detected if the following conditions were met; amplification of the kit internal control, amplification of either the *ORF1ab* or *N* gene with a cycle threshold of Cycle threshold (C_T_) < 38, detection of the positive control on the sample plate, detection of the RNA extraction control on the sample plate, and no SARS-CoV-2 specific amplification in the negative control. C_T_ values were calculated automatically using instrument software with automatic baseline setting calculated. All curves were manually inspected by two investigators in order to check for quality and inhibition of reaction.

LamPORE: Aligned read counts were generated via the LamPORE pipeline against *ORF1ab* (labelled AS1), *E1* and *N2* genes, as well as a human *ACTB* gene internal control. Any unaligned reads were marked as “Undetermined”. Samples were called positive if any of the SARS-CoV-2 target genes had > 50 reads/sample, indeterminate if between 20-50 reads and negative if less than 20 reads. ACTB gene counts were not used as part of the calling algorithm but were used to infer sufficient sampling.

Test results were compared using a 2×2 table and standard measures of sensitivity, specificity, positive predictive value and negative predictive value were calculated using R. If results from either the RT-qPCR or LamPORE test were missing or indeterminate then no comparison was made and the sample was removed from the analysis. Standard analyses of variability in diagnostic precision were made.

## RESULTS

### Participants

For the prospective asymptomatic study, a total of 1200 participants who were at work and reported to be well were recruited across the four sites (Birmingham n=600, Southampton n=200, Basingstoke/Winchester n=200, Manchester n=200). There were no adverse events. Sample flow is shown in Figure 2.

**Figure 2:**
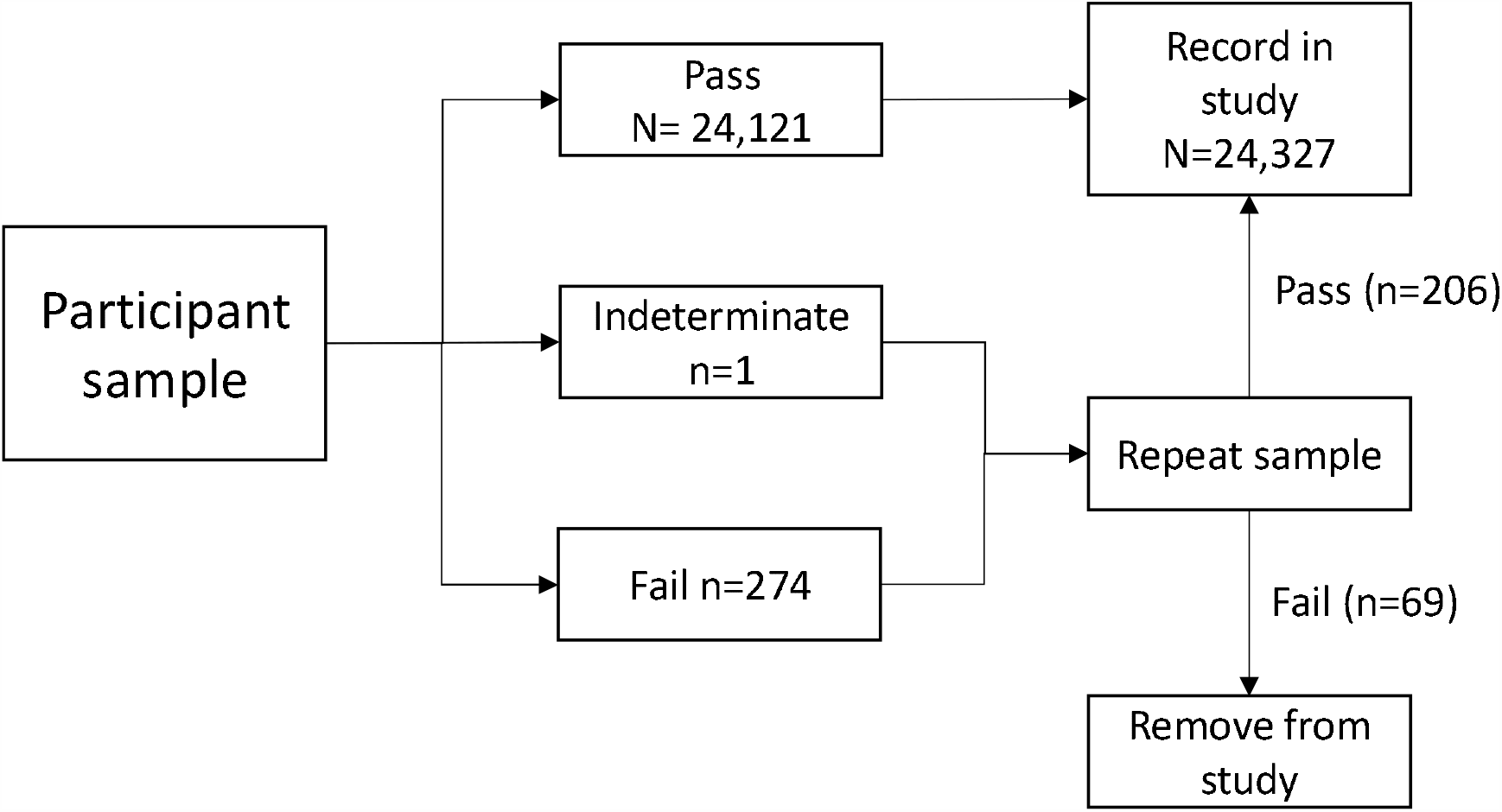
Flowchart showing sample numbers at each stage. Pass = sample passed assay QC; Indeterminate = sample passed QC but did not have a clear result; Fail = sample failed assay QC.

### Analytical performance

LamPORE reliably detected SARS-CoV-2 to 20 copies/ml of sample. SARS-CoV-2 reads were detected in the 0.2 copies/ml sample but this was below the threshold for calling as positive sample in LamPORE but were not detected via RT-qPCR (Table 1, Figure 3).

**Table 1:**
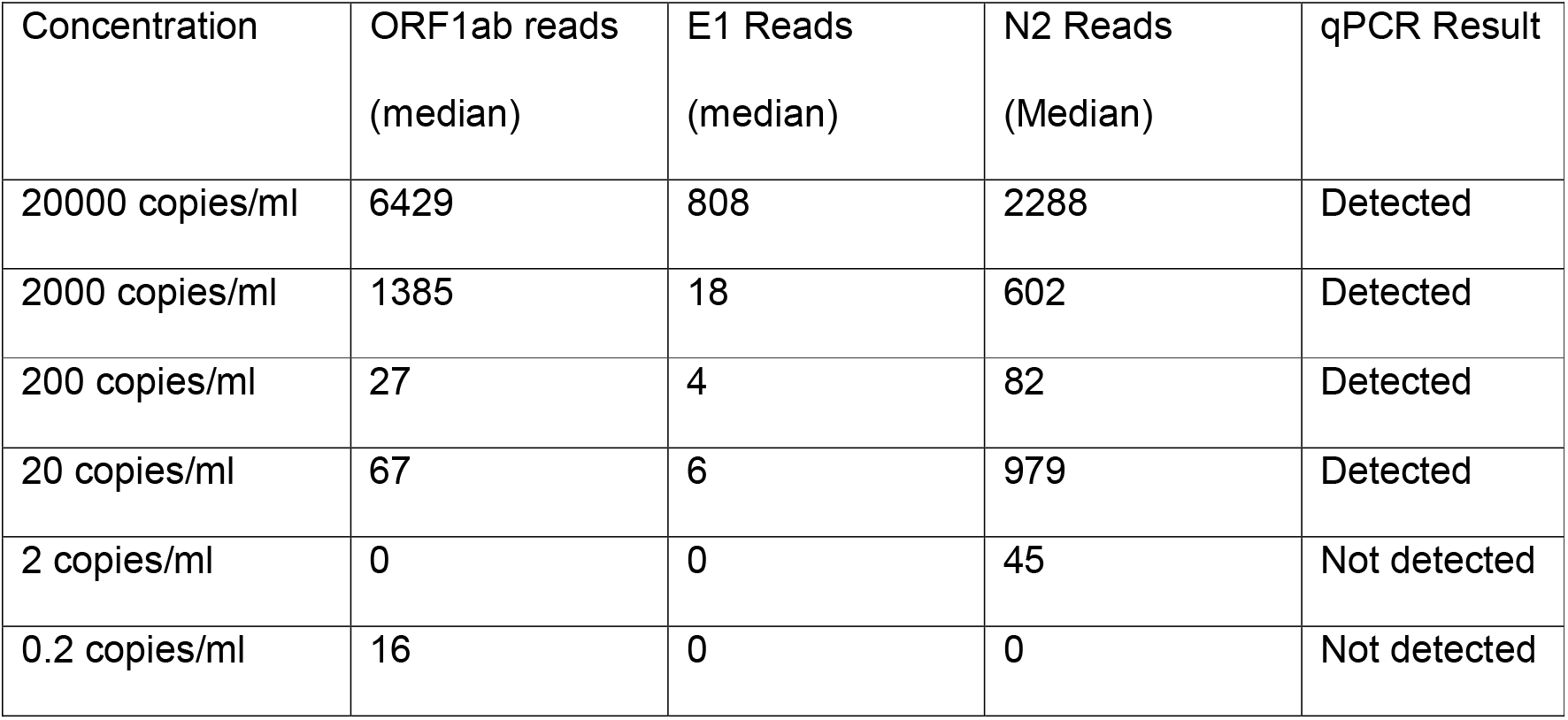
Dilution series of SARS-CoV-2 virus and LamPORE

| Concentration | ORF1ab reads<br>(median) | E1 Reads<br>(median) | N2 Reads<br>(Median) | qPCR Result |
| --- | --- | --- | --- | --- |
| 20000 copies/ml | 6429 | 808 | 2288 | Detected |
| 2000 copies/ml | 1385 | 18 | 602 | Detected |
| 200 copies/ml | 27 | 4 | 82 | Detected |
| 20 copies/ml | 67 | 6 | 979 | Detected |
| 2 copies/ml | 0 | 0 | 45 | Not detected |
| 0.2 copies/ml | 16 | 0 | 0 | Not detected |

**Figure 3:**
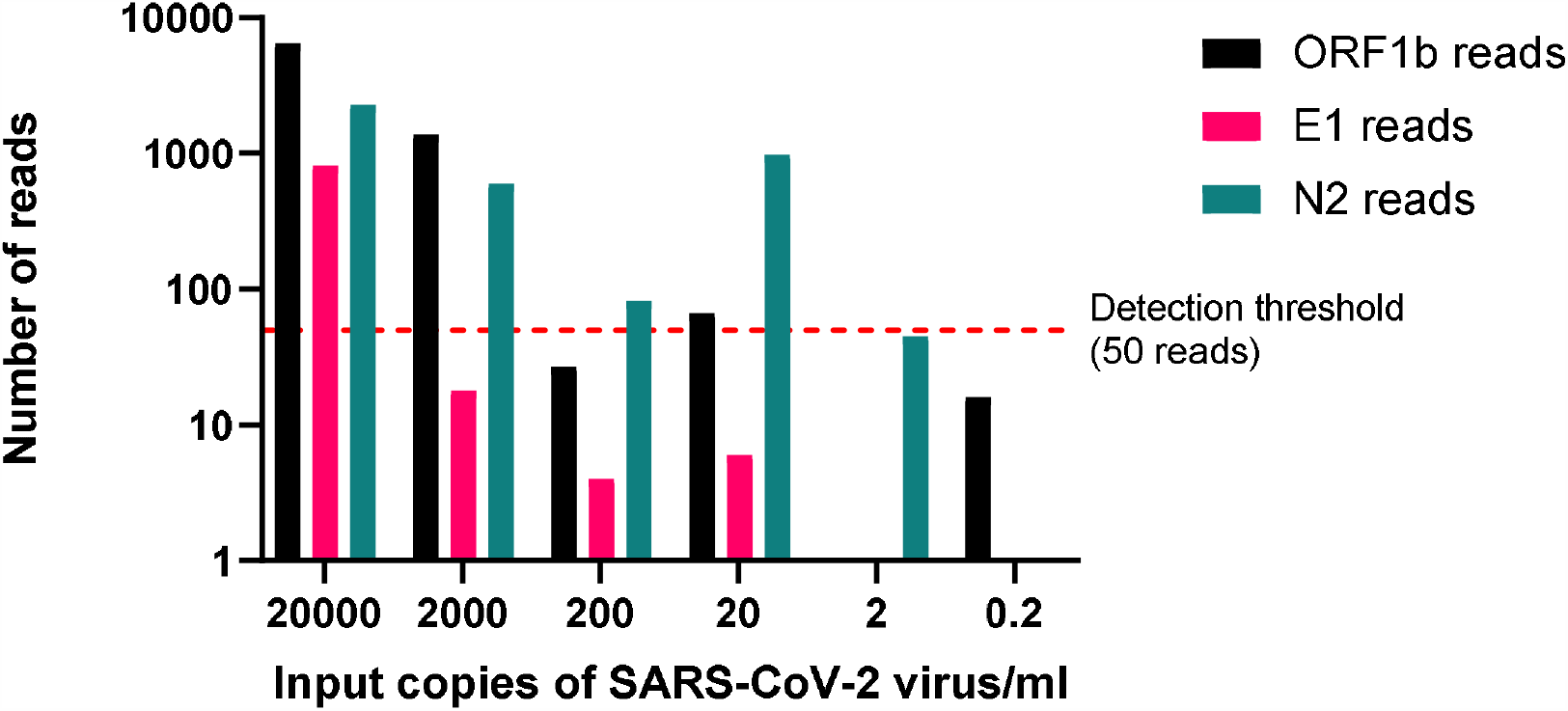
Figure showing *ORF1ab* (black), *E1* (fuchsia) and *N2* (taupe) reads in serial dilution series of SARS-CoV-2 for LamPORE. Detection threshold shown by red dotted line.

Intra- and inter-assay precision was calculated against the *ORF1ab* gene. For intra-assay precision on a single day, the standard deviation of *ORF1ab* was 50 reads with a coefficient of variation (CV) of +/- 2.3% (supplementary table 1). For inter-assay precision across multiple days, the standard deviation of *ORF1ab* was 178 reads with a CV of +/- 7.8% (Supplementary table 2).

For reproducibility for 24 replicates the standard deviation for *ORF1ab* gene was 128 reads with a CV of +/- 3.9% (Supplementary table 3).

For analytical specificity of the LamPORE assay, SARS-Cov-2 was not detected in any of the samples within the respiratory virus panel.

### Test results

#### All participants

In total 23,427 samples were obtained from all participants, of which 22,401 were from the asymptomatic study and 848 were from the retrospective symptomatic cohort. Both LamPORE (comparator assay) and RT-qPCR (reference assays) were performed on all 23,427 samples (Table 2).

**Table 2:** Diagnostic assay performance tables for RT-qPCR vs. LamPORE assay for whole cohort, asymptomatic cohort (swabs), asymptomatic cohort (saliva) and retrospective symptomatic cohort. Sensitivity, specificity, positive predictive value (PPV) and negative predictive value (NPV) for each cohort are shown at the bottom of each sub-table

| Whole cohort |  |  |  |  |
| --- | --- | --- | --- | --- |
|  |  | RT-qPCR |  |  |
|  |  | Positive | Negative |  |
| LamPORE | Positive | 477 | 124 | 601 |
|  | Negative | 2 | 22824 | 22826 |
|  |  | 479 | 22948 |  |
| Sensitivity | Specificity | PPV | NPV |  |
| 99.58 (95% CI | 99.46% (95% | 79.37% (95% | 99.99% (95% |  |
| 98.46-99.95%) | CI 99.36-99.55%) | CI 76.34-82.10%) | CI 99.9-100%) |  |
| **Asymptomatic cohort - swabs** |  |  |  |  |
|  |  | Positive | Negative |  |
| LamPORE | Positive | 23 | 11 | 34 |
|  | Negative | 0 | 3932 | 3932 |
|  |  | 23 | 3943 |  |
| Sensitivity | Specificity | PPV | NPV |  |
| 100% (95 CI 85.2-100%) | 99.72% (95% CI 99.5-99.7%) | 67.65% (95% CI 53.7-79.1%) | 100% |  |
| **Asymptomatic cohort – saliva** |  |  |  |  |
|  |  | Positive | Negative |  |
| LamPORE | Positive | 187 | 112 | 299 |
|  | Negative | 2 | 18134 | 18136 |
|  |  | 189 | 18246 |  |
| Sensitivity | Specificity | PPV | NPV |  |
| 98.9% (95% CI 96.2-99.9%) | 99.4% (95% CI 99.3-99.5%) | 62.5% (95% CI 58.1-66.8%) | 99.9% (95% CI 99.9-100%) |  |
| **Symptomatic cohort** |  |  |  |  |
|  |  | Positive | Negative |  |
| LamPORE | Positive | 116 | 0 | 116 |
|  | Negative | 0 | 752 | 752 |
|  |  | 116 | 752 | 868 |
| Sensitivity | Specificity | PPV | NPV |  |
| 100% (95% CI<br>96.9%-100%) | 100% (95% CI<br>99.51-100%) | 100% | 100% |  |

Of the 601 samples positive on LamPORE assay, 477 were also positive and 124 negative by RT-qPCR and comparison to the reference assay and 124 were false positives. Of the 22,826 negative samples, 22,824 were confirmed as negative by the RT-qPCR and there were two samples that were positive by RT-qPCR.

The diagnostic sensitivity (DSe) of the LamPORE assay compared to the Certest ViaSure RT-qPCR assay was 99.58% (95% CI 98.46-99.95%) and the diagnostic specificity (DSp) was 99.46% (95% CI 99.36-99.55%). The positive predictive value (PPV) of the test with a tested population incidence of 2.04% was 79.37% (95% CI 76.34-82.10%) and the negative predictive value calculated with a prevalence of 2.04% (NPV) was 99.99% (95% CI 99.9-100.0%).

When modelled at 1% population prevalence the PPV dropped to 66.24% and at 0.1% population prevalence the PPV was 16.3%. NPV remained at > 99.99% in all population scenarios.

#### Asymptomatic cohort

For the asymptomatic cohort (table 2) a total of 22,401 participant samples were tested, with 333 positive (34 swab, 299 saliva) samples being identified of which 210 samples (23 swab, 187 saliva) were true positive and 123 samples (11 swab, 112 saliva) false positive when compared with RT-qPCR. There were 22,068 negative samples in total of which 22,026 samples (3932 swab, 18134 saliva) were true negative and 2 samples (both saliva) false negatives. For this cohort there was a diagnostic sensitivity (DSe) of 99.64% (95% CI 98.0-99.9%), a diagnostic specificity (DSp) of 99.48% (95% CI 99.38-99.57%), a PPV of 69.44% and an NPV of 99.48% (95% CI 99.97-100.00%). For the RT-qPCR assay, the mean *ORF1ab* cycle threshold (Ct) was 17.1 (Range 16.2-37.2) and the *N1* was 14.3 (range 11.0-37.2).

#### Symptomatic cohort

There was complete agreement between the RT-qPCR and LamPORE assays for 116 positive samples and 752 negative samples, for this cohort resulting in a DSe of 100%, DSp of 100%, PPV of 100% and NPV of 100%. The incidence of SARS-CoV-2 was 13.4% over the study period.

### Variability across time course

In order to understand the utility of LamPORE across the time course of infection, a single participant who was identified as the beginning of their infection, early in the study with a long time course (5 days) of positivity was studied with daily saliva sampling as per protocol (Figure 4). Initially a high viral load, indicative of a C_T_ value of 19.5 was observed which increased (indicating decreasing viral load) over the five days to 23.5 and then became undetectable at day 6. Only LamPORE *N2* reads were detectable at day 1, but *E1* reads became detectable at day 2 and *ORF1ab* reads at day 4.

**Figure 4:**
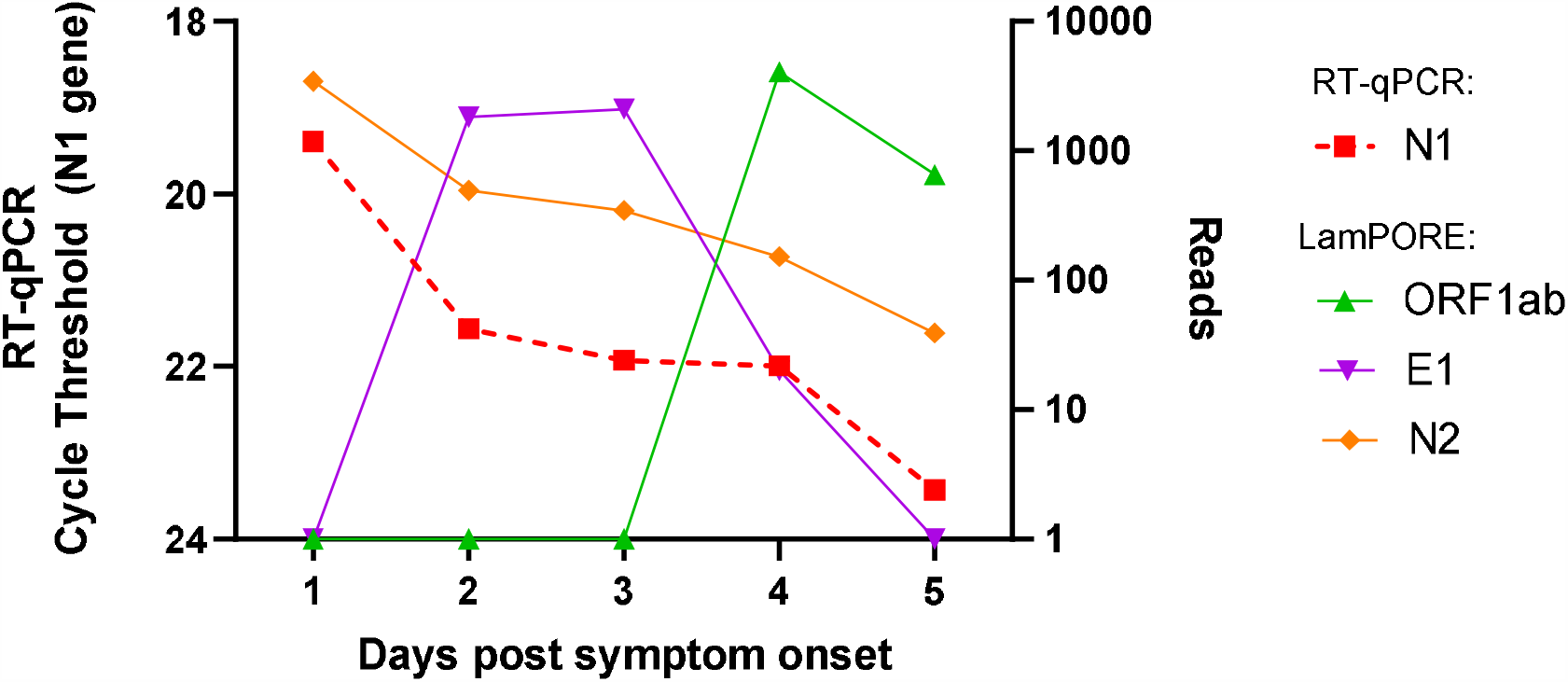
Line plot showing data from daily saliva sampling of a single participant reporting symptoms and their cycle threshold for the *N1* gene (red dashed line, left Y axis, reverse order) and read count (right Y axis) for *ORF1ab* (green line), *E1* (purple line) and *N2* (orange line). Days since symptoms began shown on X axis.

## DISCUSSION

We carried out a very large asymptomatic cohort study of health care workers using a novel technology, LamPORE, comparing it to a reference CE-IVD marked RT-qPCR assay. In addition, we have also carried out a large symptomatic cohort study based on “real world” samples typically found in clinical practice. We found that LamPORE has high sensitivity and specificity (>99%) in both the asymptomatic and symptomatic populations, directly comparable to RT-qPCR and therefore has comparable predictive ability across a range of use cases in varying levels of population prevalence. We studied a population with a wide range of viral loads as determined by cycle threshold, with LamPORE demonstrating good detection across the range, suggesting it has applicability across the whole spectrum of use cases.

LamPORE has the advantage that it is scalable (11) to allow testing of very large population levels because of the use of sample barcoding allowing pooling of up to 3,500 samples on a single GridIon instrument. Due to the increased sensitivity of LAMP as part of the LamPORE system, it gives very high sensitivity for SARS-CoV-2. LamPORE has the ability to be automated, reducing error rates and increasing throughput. With current LamPORE barcoding capability 768 samples can be sequenced simultaneously on one MinIon flow cell. With combinatorial barcoding as has been adopted in other population level assays (15) on a larger flow cells (e.g. a Promethion flow cell), even greater sample multiplexing may be achievable. Another potential inherent advantage is the ability to multiplex gene targets allowing the detection of multiple respiratory pathogens (16) such as SARS-CoV-2, influenza and respiratory syncytial virus (RSV). It is not known what the upper limit of multiplexing of LAMP primers is, and they are considerably more complex to design than PCR primers (17). Given the advantages of LAMP in terms of speed of amplification (9) and sensitivity of detection, an exploration of LAMP multiplexing is urgently required. Also, the assay chemistry uses different enzymes and methodologies to PCR, meaning a diversification of supplies and therefore potentially less reagent shortages in a pandemic setting.

A potential disadvantage of LamPORE is the differing workflow needed to prepare samples, including the LAMP step and library preparation, barcoding then sequencing. This requires more sample preparation steps than a RT-qPCR workflow. RNA extraction remains the real rate limiting step, for both LamPORE and RT-qPCR and ourselves and other groups are currently working on direct alternatives removing the need for RNA extraction.

During the testing of the asymptomatic cohort, we observed a number of false positives using LamPORE when compared to the RT-qPCR assay. There are a number of possible explanations for this observation. Firstly, LAMP amplification is more sensitive than PCR amplification (18) and so contamination risk is high but as the laboratories refined the technique, contamination issues seemed to resolve. Secondly, it is feasible that some of the samples are in fact, true positive as demonstrated by the ability of LamPORE to detect spiked, killed virus beyond the limit of detection of RT-qPCR. This may have useful implications for sample pooling (19), as greater sensitivity would allow more samples to be pooled and tested. Finally, the LamPORE protocol requires multiple manual liquid handling steps which can lead to error and increases the number of opportunities for contamination to occur. The use of automated liquid handling methods (as developed by the manufacturer) is likely to reduce or eliminate this phenomenon.

In conclusion, we have demonstrated the accuracy of LamPORE across a range of population use cases, maintaining a high specificity and sensitivity, reproducibility and limit of detection, as well as working well on saliva samples, making it suitable for the detection of symptomatic and asymptomatic patients with SARS-CoV-2.

## Supporting information

STARD Checklist LamPORE

## Data Availability

Data available on reasonable request and after central review

## SUPPLEMENTARY MATERIALS

**Supplementary table 1:**
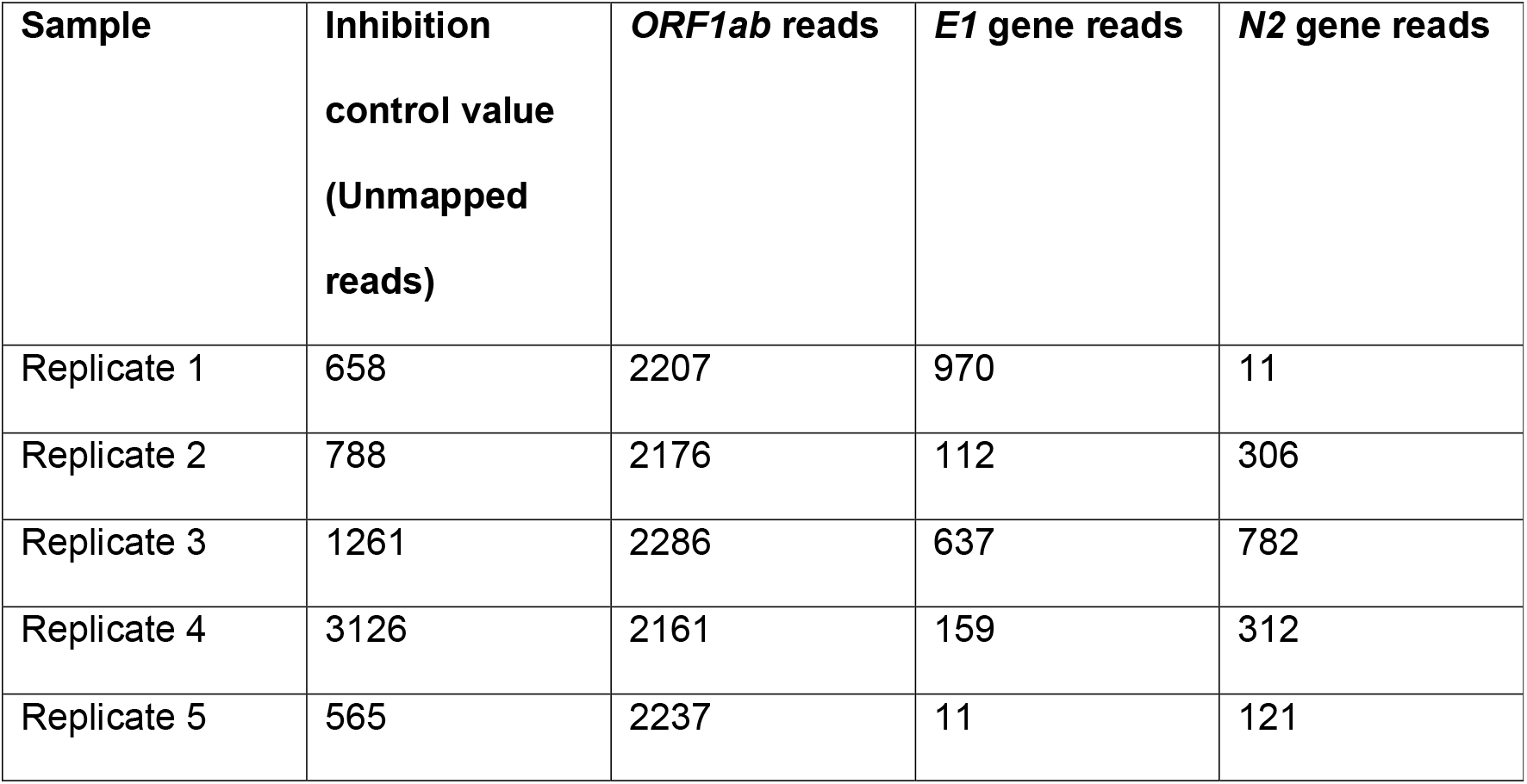
Intra-assay precision. Standard deviation (SD) *ORF1ab* = 50 reads, coefficient of variation (CV) = +/- 2.3%

**Supplementary table 2:**
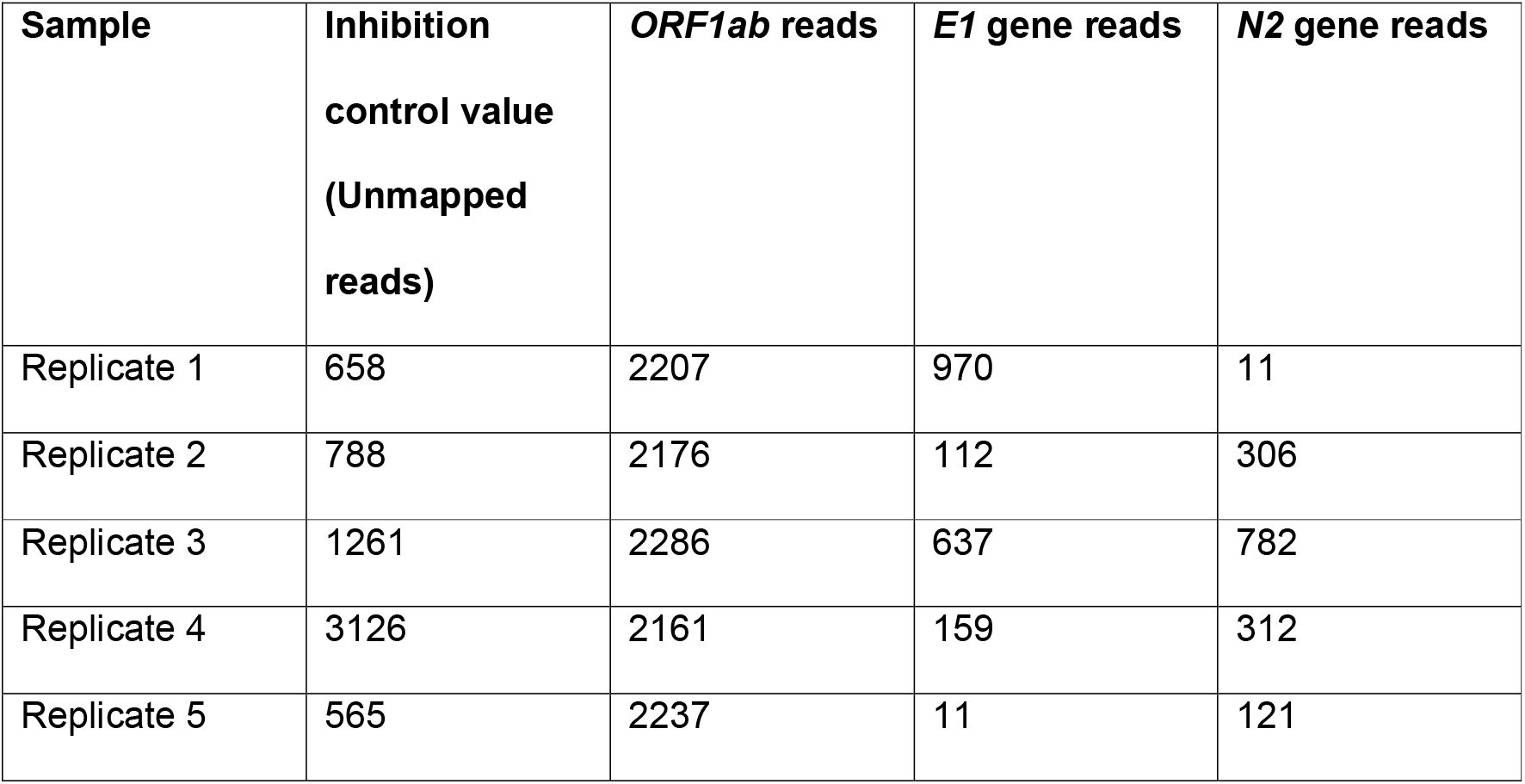
Inter-assay precision. Standard deviation (SD) *ORF1ab* = 178 reads, CV = +/- 7.8%

**Supplementary table 3:**
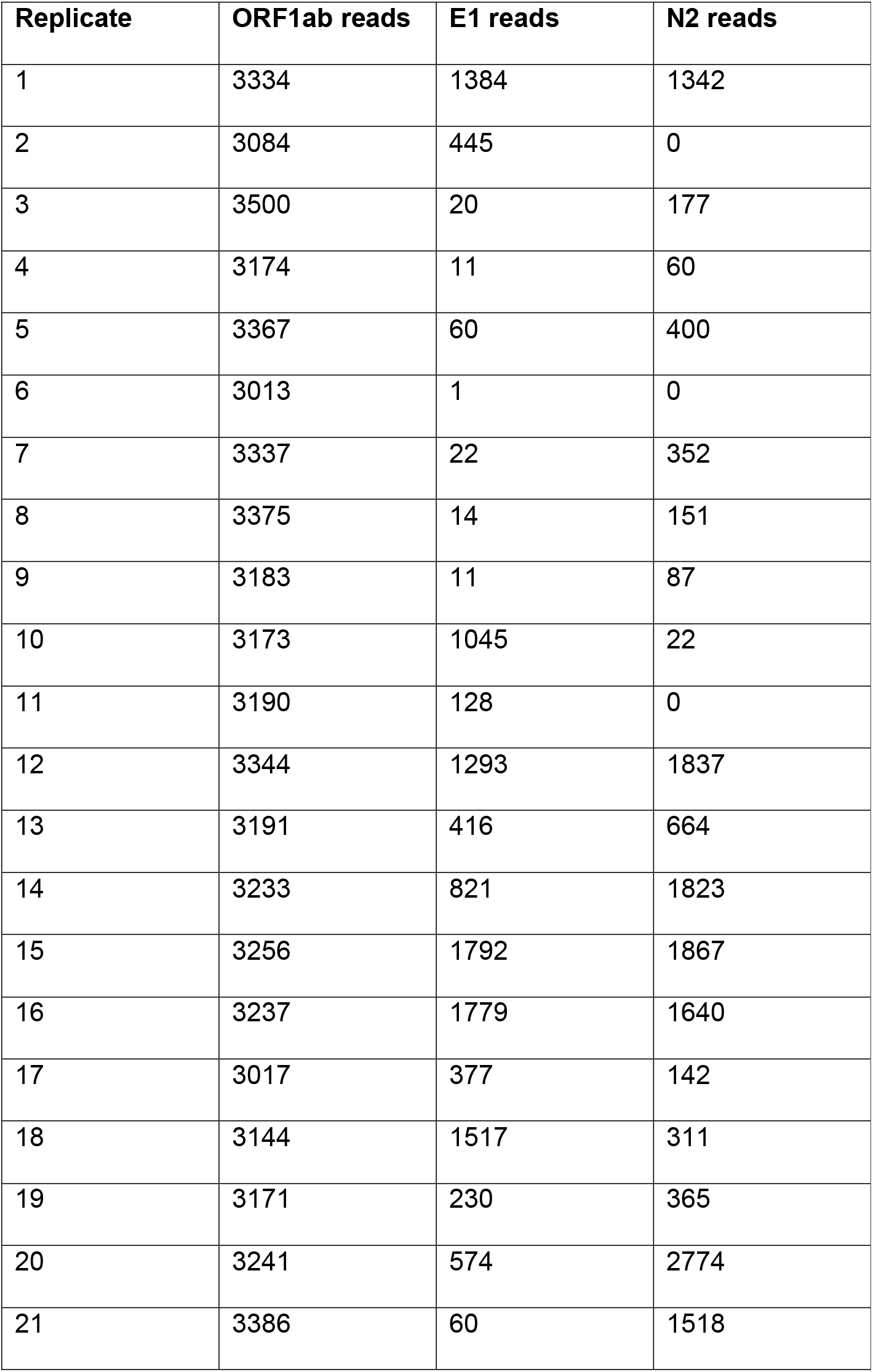
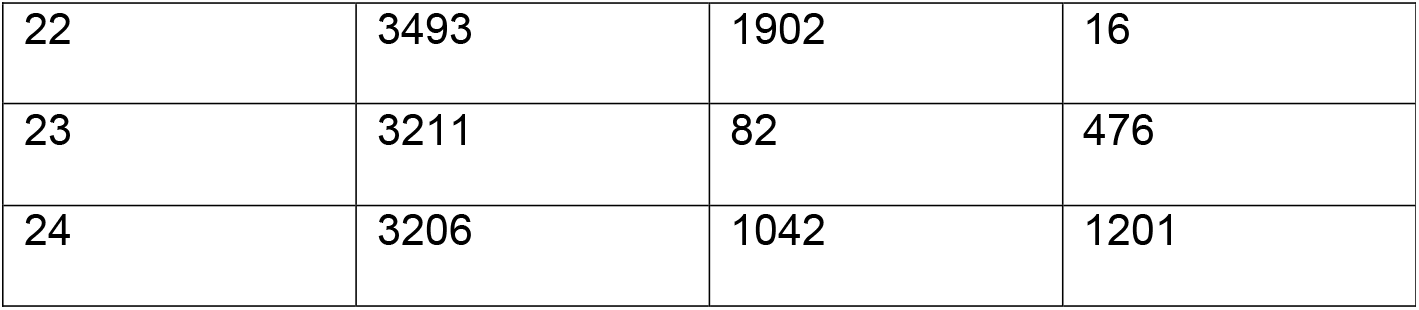
Assay variability. Standard deviation (SD) ORF1ab = 128 reads with CV = +/- 3.9%

